# MRI-Derived Variables Combined with Machine Learning for Pulmonary Hypertension Risk Prediction: A Retrospective Analysis

**DOI:** 10.1101/2025.02.28.25323129

**Authors:** Hongxia Jiang, Fan Yang, Changrong Wu, Dan Xiao, Xiaojun Hao, Zhenshun Cheng

**Author notes:** Corresponding author: Zhenshun Cheng, Address: No.169, Donghu Road, Wuchang District, Wuhan City, Hubei Province, P.R. China. These authors contribute equally to this work (Hongxia Jiang, Fan Yang, Changrong Wu, Dan Xiao).

## Abstract

**Background:** Pulmonary hypertension (PH) is a severe and progressive vascular disease for which early diagnosis and risk stratification are critical for improving patient outcomes. However, current diagnostic approaches exhibit limitations in achieving precise risk prediction. This study aimed to develop and validate a machine learning-based risk prediction model that integrates MRI-derived variables for PH risk assessment.

**Methods:** This retrospective study enrolled 210 participants who underwent MRI at Zhongnan Hospital of Wuhan University between January 2021 and December 2023, including 87 PH patients and 123 controls. Key MRI features were selected through recursive feature elimination (RFE) with a Random Forest algorithm. Multiple machine learning models, including XGBoost and logistic regression, were trained and evaluated employing 10-fold cross-validation. Model performance was evaluated using receiver operating characteristic (ROC) curves, calibration curves, and decision curve analysis (DCA). SHAP analysis was utilized to interpret the contribution of individual features, and a nomogram integrating MRI and clinical variables was developed for personalized risk prediction.

**Results:** Six key MRI-derived features were identified, among which pulmonary artery diameter and left atrial anterior-posterior diameter were the most significant predictors. The XGBoost model exhibited the best performance, achieving an area under the curve (AUC) values of 1.0 and 0.969 in the training and testing sets, respectively. Calibration curves demonstrated excellent agreement between predicted and observed outcomes. DCA revealed high net clinical benefits across a range of risk thresholds. The developed nomogram offers an intuitive tool for individualized PH risk prediction, demonstrating strong interpretability and clinical utility.

**Conclusion:** This study developed a highly accurate and reliable machine learning-based risk prediction model for PH based on MRI-derived features. By integrating SHAP analysis and a nomogram, the model provides a novel, non-invasive approach for early diagnosis and personalized risk stratification of PH, highlighting the significant potential for clinical application.

## Introduction

Pulmonary hypertension (PH) is a severe, progressive vascular disorder characterized by elevated pulmonary arterial pressure, increased pulmonary vascular resistance, and eventual right heart failure [1]. Epidemiologically, PH affects approximately 1% of the global population, with its prevalence increasing to 10% among individuals aged over 65 years. The disease imposes a substantial burden on patients, resulting in high morbidity, reduced quality of life, and significant mortality [2]. Despite therapeutic advancements, PH remains a life-threatening condition, with untreated patients exhibiting a median survival of less than three years [3]. Early detection and accurate risk stratification are critical for improving clinical outcomes, facilitating timely therapeutic interventions, and mitigating the disease’s impact.

Current diagnostic strategies for PH encompass both invasive and non-invasive modalities. Right heart catheterization (RHC) is regarded as the gold standard for confirming PH diagnosis and assessing hemodynamic parameters [4]. However, its invasive nature, associated risks, and limited accessibility render it unsuitable for routine screening or monitoring. Echocardiography is widely utilized as a non-invasive screening tool, offering estimates of pulmonary artery pressure and insights into right ventricular function [5]. Although valuable, echocardiography is operator-dependent and exhibits insufficient sensitivity and specificity for reliably detecting early-stage PH.

Magnetic resonance imaging (MRI) has emerged as a highly accurate and non-invasive modality for assessing cardiac and pulmonary vascular structures [6]. It provides detailed and reproducible assessments of cardiovascular morphology and function, rendering it particularly valuable for elucidating the structural and hemodynamic changes associated with pulmonary hypertension (PH). In addition to imaging, laboratory biomarkers offer complementary insights into the biochemical and physiological alterations underlying the disease. Integrating imaging-derived and biochemical data enhances diagnostic precision by combining structural and functional aspects of PH, facilitating a more comprehensive evaluation.

Despite the abundance of imaging and laboratory data, integrating multidimensional datasets into a unified predictive framework remains a significant challenge. Advances in bioinformatics and machine learning offer powerful tools for addressing this complexity [7]. These approaches facilitate the selection of key features, optimization of predictive algorithms, and development of interpretable models capable of stratifying PH risk with high accuracy. By leveraging MRI and laboratory data, machine learning models can identify high-risk individuals, facilitate early diagnosis, and guide personalized treatment decisions.

In this study, we aim to develop and validate a machine learning-based predictive model that integrates MRI-derived indices and laboratory biomarkers for PH risk stratification. By integrating advanced imaging, laboratory data, and bioinformatics techniques, we propose a novel non-invasive tool for early PH detection and individualized patient management, addressing critical gaps in current diagnostic and prognostic approaches.

## Materials and Methods

### Study Design and Population

This retrospective study enrolled data from 210 individuals, comprising 87 patients with PH and 123 control subjects (Figure 1). All participants underwent MRI and clinical evaluations at Zhongnan Hospital of Wuhan University from January 2021 to December 2023. The PH group comprised individuals diagnosed with PH through comprehensive clinical assessments and imaging studies, with the diagnosis confirmed by right heart catheterization (RHC) showing elevated pulmonary arterial pressure. Participants were included if they were aged over 14 years. Patients were excluded if their mean pulmonary arterial pressure (mPAP) measured by RHC was below 20 mmHg or if RHC data were incomplete or unavailable.

**Figure 1.**
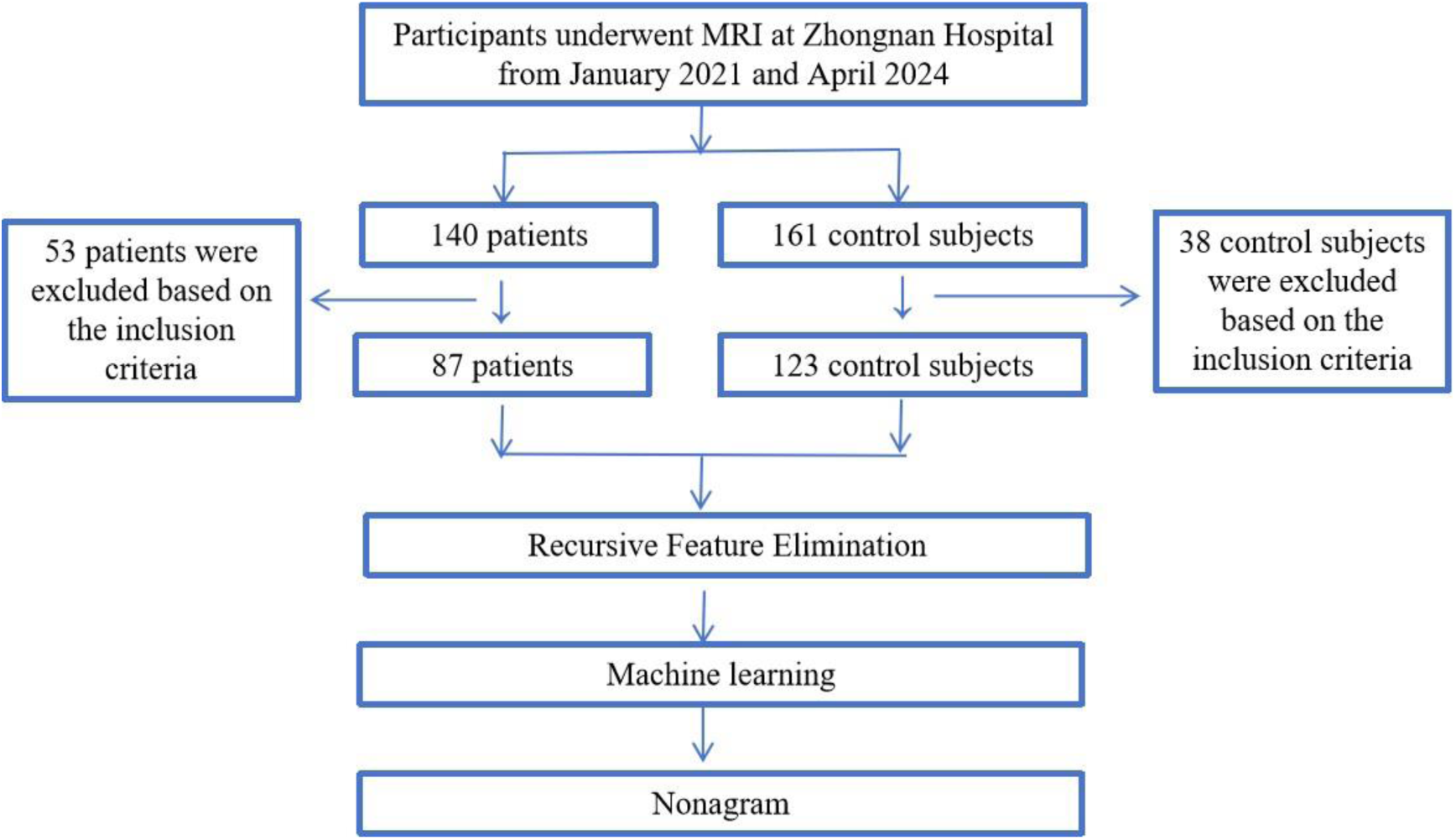
Flowchat of the study

Control subjects were selected based on the absence of significant abnormalities in cardiac structure, valve function, or ventricular wall motion as assessed by MRI. All participants in the control group were also required to be aged over 14 years. To ensure data integrity, individuals with incomplete datasets, defined as those with more than 5% missing data, were excluded from the study. This study was approved by the Ethics Committee of Zhongnan Hospital of Wuhan University (Approval No. 2023185). All patient data were anonymized to ensure confidentiality.

### Data Collection and Variables

This study retrospectively collected data from electronic medical records at Zhongnan Hospital of Wuhan University between January 2021 and December 2023. The dataset included demographic information, imaging parameters, and laboratory test results.

Demographic data, including age and gender, were recorded for all participants. Imaging parameters assessed cardiac structure and function using magnetic resonance imaging (MRI). Key measurements included pulmonary artery dimensions, atrial and ventricular sizes, interventricular septal thickness, and aortic diameter. Functional parameters, such as flow velocity, regurgitation severity, and pressure gradients for cardiac valves, were also obtained. All imaging measurements were verified by two independent radiologists to ensure accuracy. Laboratory test results included complete blood count, hematological parameters, coagulation profiles, liver and kidney function tests, electrolytes, lipid profiles, thyroid function, immune markers, and arterial blood gas analysis. These variables provided additional insights into the physiological and biochemical alterations associated with pulmonary hypertension.

### Data Preprocessing

In the data preprocessing step, R software (Version 4.4.1) and relevant packages were used to prepare the dataset for analysis. Missing values were handled using median imputation, where missing entries were replaced with the median value of the corresponding variable to maintain data completeness. Continuous variables were centered and standardized to have a mean of 0 and a standard deviation of 1, ensuring compatibility with machine learning algorithms. Categorical variables were encoded as factors to ensure accuracy in subsequent statistical and computational analyses. These preprocessing steps ensured that the dataset was consistent, complete, and ready for feature selection and model development.

### Feature Selection and Model Development

We used stratified sampling to randomly split the dataset into training and testing sets to ensure a balanced representation of PH and control groups. For feature selection from MRI-derived variables, Recursive Feature Elimination (RFE) was applied using a Random Forest-based approach. RFE iteratively removed features contributing the least to model performance, identifying an optimal subset of variables. To ensure robustness and generalizability, we employed 10-fold cross-validation during the feature selection process.

Using the selected MRI-derived features, multiple machine learning models were trained and evaluated, including logistic regression, elastic net, decision tree, random forest, XGBoost, support vector machine (SVM), k-nearest neighbors (KNN), naive Bayes, and gradient boosting machine (GBM). Model training and evaluation were conducted primarily using the caret R package. Hyperparameter optimization was performed using grid search combined with 10-fold cross-validation. The Area Under the Receiver Operating Characteristic Curve (AUC-ROC) was used as the primary performance metric and was computed and visualized using the pROC R package. The model with the highest AUC-ROC was selected as the optimal model for predicting PH based on MRI-derived variables.

For internal validation, bootstrap resampling was applied to evaluate model stability and performance. To enhance model interpretability, Shapley Additive Explanations (SHAP) were used to assess the contribution of individual variables to the model’s predictions. These insights allow for a better understanding of feature importance and their relationship to PH risk.

The final predictive model was developed by combining the selected MRI-derived and clinical features into a comprehensive framework. Model performance was evaluated on the test dataset, and interpretability was further enhanced through SHAP analysis. The results provided a robust, interpretable, and clinically applicable tool for PH risk stratification.

### Nomogram Construction

A nomogram was constructed using the rms package in R, based on the optimal logistic regression model. The nomogram translates the contributions of each predictive variable into a total score, which corresponds to the probability of developing PH. This tool provides an intuitive and practical way for clinicians to assess individual risk. Calibration curves confirmed the alignment between predicted and observed probabilities, ensuring reliability and clinical utility.

### Evaluation Metrics

To assess the performance of the predictive model and nomogram, the Receiver Operating Characteristic (ROC) curve was used as the primary evaluation metric, measuring the model’s ability to discriminate between PH patients and controls. The Area Under the Curve (AUC) was calculated to quantify model performance.

Calibration curves were plotted to evaluate the agreement between predicted probabilities and observed outcomes. These curves ensured the reliability of the model’s predictions.

Decision curve analysis (DCA) was performed using the rmda package to quantify the net clinical benefit across varying threshold probabilities. This analysis highlighted the model’s utility in clinical decision-making compared to alternative strategies.

Finally, an interactive online tool was developed using the shiny R package (version 1.9.1) to facilitate the application of the predictive model in clinical practice, enabling real-time risk assessment for PH.

### Statistical Analysis

Feature contribution analysis was conducted using SHAP (Version 0.46.0) in Python (Version 3.10.8) to interpret the impact of individual variables on the predictive model. All other statistical computations were performed in R (Version 4.4.1).

To determine the distribution of continuous variables, the Shapiro-Wilk test was applied. Variables exhibiting normal distributions were compared using independent-sample t-tests. For non-normally distributed data, non-parametric methods such as the Mann-Whitney U test or the Kruskal-Wallis test were employed based on the number of groups. Categorical variables were summarized as percentages and evaluated for group differences using either the Chi-square test or Fisher’s exact test for smaller sample sizes. A significance level of P < 0.05 was used for all tests. These methods ensured a comprehensive and accurate statistical assessment of group differences and model performance.

## RESULTS

### Comparison of MRI Parameters and Laboratory Findings between Pulmonary Hypertension Patients and Healthy Controls

This study enrolled 210 participants, including 123 controls and 87 patients with PH. Significant differences were observed between the PH and control groups in multiple MRI parameters and laboratory findings. PH patients exhibited larger dimensions in the left atrium (LA), left ventricle (LV), right atrium (RA), and right ventricle (RV), along with reduced left ventricular ejection fraction (LVEF) and impaired ventricular systolic and diastolic function (all P < 0.05). Hematological tests showed significantly higher neutrophil percentages and lower lymphocyte percentages in PH patients compared to controls (P < 0.05). Coagulation function tests indicated prolonged prothrombin time (PT) and elevated international normalized ratio (INR) in PH patients (P < 0.05). Furthermore, liver function markers, including total bilirubin, direct bilirubin, and indirect bilirubin, were significantly elevated in PH patients (P < 0.05). Electrolyte analysis showed reduced levels of serum potassium, sodium, and phosphorus, whereas calcium and chloride levels were elevated in PH patients (P < 0.05) (Table).

### Identifying Key Features and Evaluating Model Performance for Pulmonary Hypertension Prediction

This study employed RFE combined with a Random Forest model to identify the most significant MRI-based predictors for PH. A total of 6 key features were selected through 10-fold cross-validation, with pulmonary artery (PA) diameter and left atrial anterior and posterior diameter emerging as the most impactful predictors (Figure 2A). The relationship between the number of selected features and model accuracy during the RFE process is illustrated in Figure 2B, where the model’s performance plateaus when 10 to 16 features are included, marking this range as optimal for prediction.

**Figure 2.**
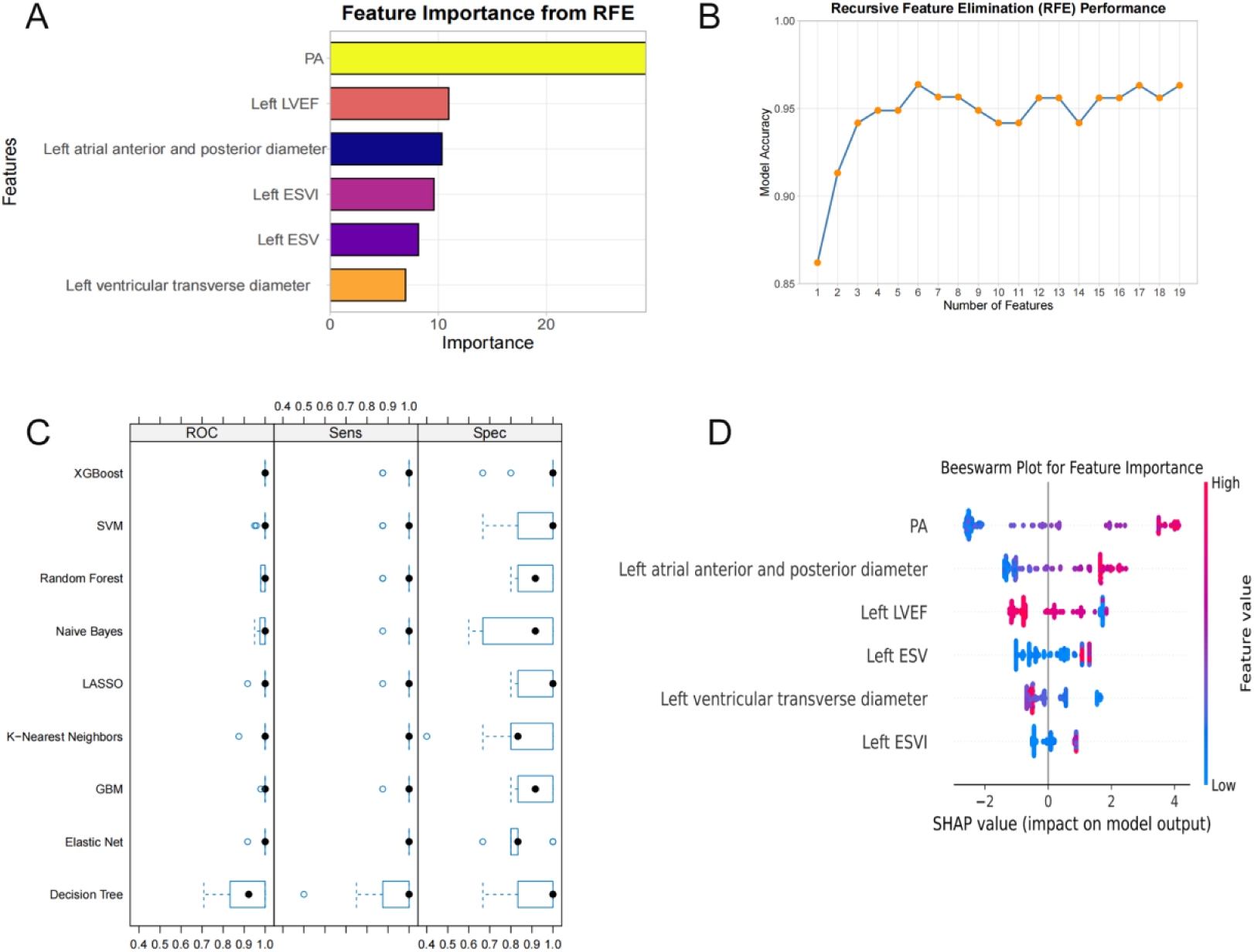
Feature selection, model performance, and interpretability for pulmonary hypertension prediction. (A)Feature importance identified by RFE; (B)Recursive Feature Elimination performance; (C)Machine learning model performance comparison; (D)SHAP analysis for model interpretability.

Among the evaluated models, XGBoost demonstrated the best overall performance, achieving high scores in AUC, sensitivity, and specificity (Figure 2C), reflecting its robustness in minimizing misclassifications. SHAP analysis (Figure 2D) highlighted PA diameter, left atrial anterior and posterior diameter, and left ventricular ejection fraction as the most impactful features, confirming their key role in distinguishing PH patients from controls.

A training-to-validation split ratio of 0.65 was employed to validate the model. The confusion matrix for the initial evaluation (Figure 3A) indicated strong predictive accuracy for the control group but revealed a notable number of false positives for the PH group. A refined model (Figure 3F) demonstrated a reduction in false positives, resulting in improved predictive balance and accuracy for both classes.

**Figure 3:**
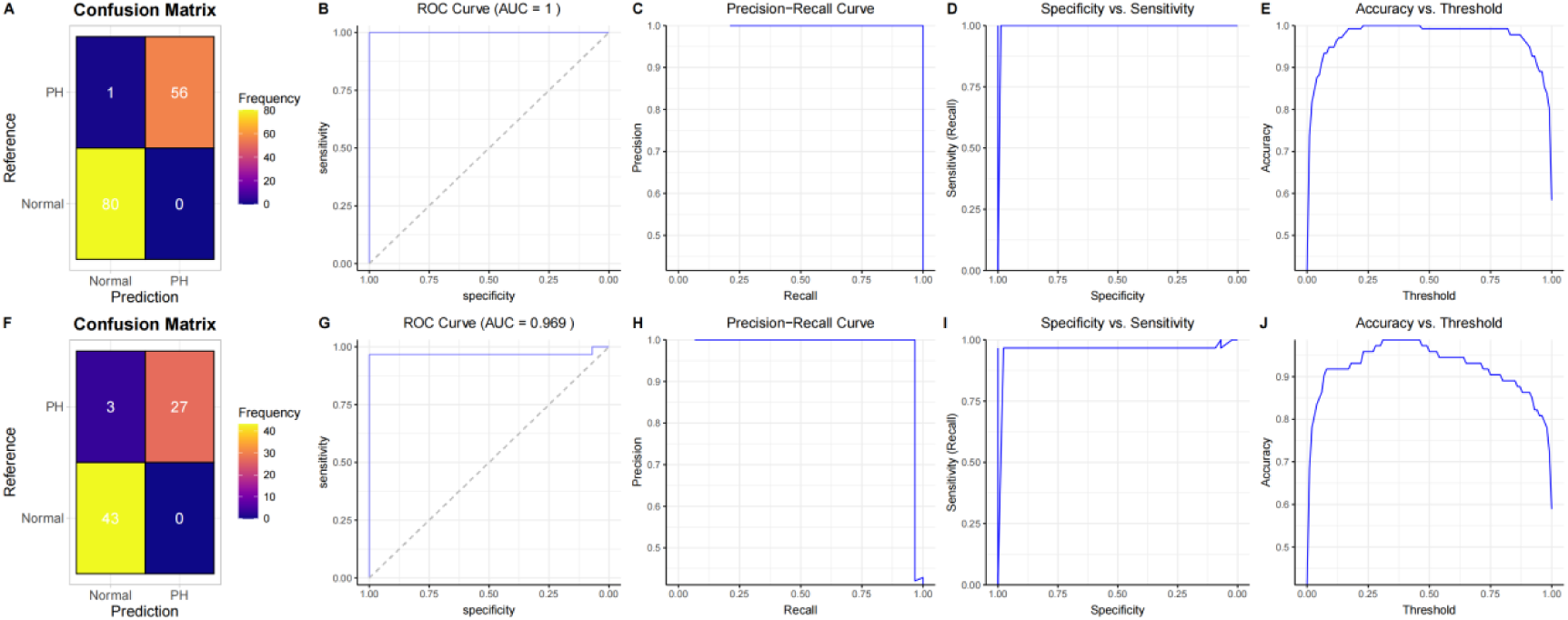
Model evaluation and refinement for pulmonary hypertension prediction. (A) Confusion matrix for the initial model; (B) Receiver Operating Characteristic (ROC) curve for the initial model; (C) Precision-Recall (PR) curve for the initial model; (D) Sensitivity and specificity balance across decision thresholds for the initial model; (E) Accuracy vs. threshold curve for the initial model; (F) Confusion matrix for the refined model; (G) ROC curve for the refined model; (H) PR curve for the refined model; (I) Sensitivity and specificity balance for the refined model; (J) Accuracy vs. threshold curve for the refined model.

The ROC curves (Figures 3B and 3G) indicated excellent discriminative ability, with The AUC values in the training and testing sets being 1 and 0.969, confirming the model’s robustness in classifying PH and control groups. Precision-recall curves (Figures 3C and 3H) further validated the model’s high performance, with precision values remaining near 1 for a wide range of recall values, demonstrating its ability to accurately identify true positives while minimizing false positives.

Specificity and sensitivity metrics (Figures 3D and 3I) were both near 1, achieving an effective balance between reducing false positives and false negatives. Accuracy vs. Threshold curves (Figures 3E and 3J) showed stable performance, with optimal accuracy achieved at thresholds ranging from 0.7 to 0.8, highlighting the model’s reliability across varying decision thresholds.

Overall, the XGBoost model demonstrated outstanding predictive performance with high precision, sensitivity, and specificity, making it a robust and reliable tool for PH diagnosis. The combination of feature selection and advanced machine learning techniques ensures that the model not only performs well but also provides interpretable insights into the factors critical for PH prediction.

### Development and Validation of a Predictive Model for Pulmonary Hypertension

To predict the risk of pulmonary hypertension (PH), a logistic regression model was constructed, and a visually intuitive nomogram was developed. The nomogram provides an individualized scoring system for each predictor variable, allowing for the transformation of aggregate scores into corresponding PH risk probabilities using a linear formula (Figure 4A).

**Figure 4.**
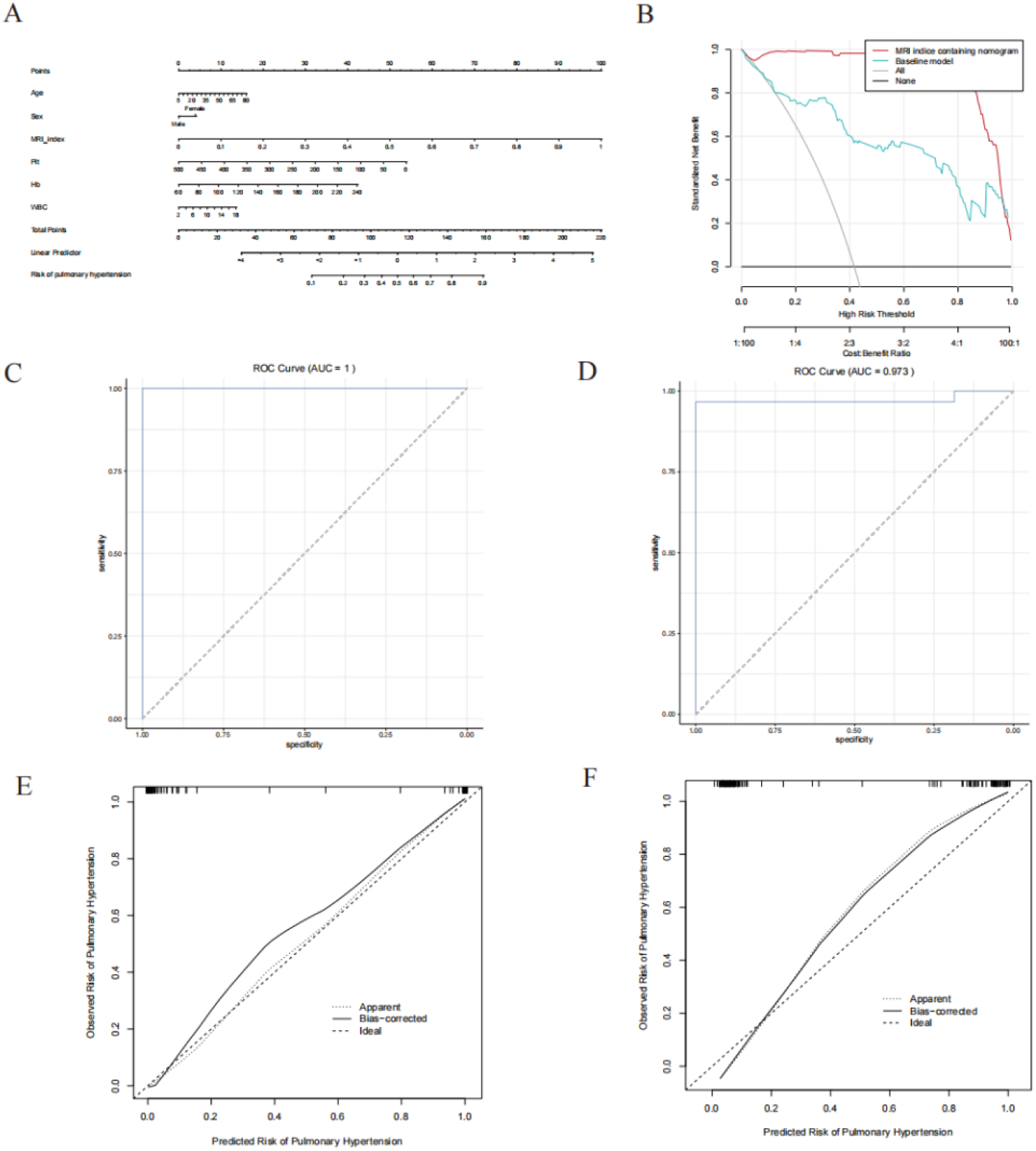
Nomogram construction and clinical utility for pulmonary hypertension risk prediction. (A)Nomogram for Risk Prediction of Pulmonary Hypertension; (B)Decision Curve Analysis;(C) ROC curve for the training sets; (D) ROC curve for the testing sets; (E)Calibration curve of the nomogram in the developing cohort. (F) Calibration curve of the nomogram in the validation cohort.

Decision Curve Analysis (DCA) highlighted the clinical utility of the nomogram. The analysis demonstrated that the nomogram consistently outperformed alternative approaches, providing a greater net clinical benefit across a range of threshold probabilities, particularly at higher PH risk thresholds (Figure 4B).

Receiver Operating Characteristic (ROC) curves were used to evaluate the discriminative ability of the model. In the test set, the initial model achieved a satisfactory area under the curve (AUC-ROC=1) (Figure 4C). However, in the validation set, the AUC value slightly decreased(AUC-ROC=0.973) (Figure 4D), indicating some performance variation during external validation, possibly reflecting slight overfitting.

Calibration curves further validated the accuracy of the predictions, showing strong agreement between predicted probabilities and actual outcomes (Figure 4E and F). This indicates a high degree of calibration and reliability.

These findings collectively indicate that the nomogram is a reliable and effective tool for predicting pulmonary hypertension risk. Even with minor decreases in AUC values, it retains strong calibration, discrimination, and clinical applicability.

### Integrated MRI and Clinical Variable Tools for Pulmonary Hypertension Risk Prediction

Figure 5A presents a comprehensive risk assessment tool for pulmonary hypertension (PH) utilizing MRI-derived parameters such as PA diameter, left atrial anterior and posterior diameter, and left ventricular ejection fraction to calculate a risk index. SHAP analysis displayed alongside the tool provides an intuitive explanation of the importance of each variable in the prediction, allowing clinicians to assess patient risk effectively using MRI data.

**Figure 5.**
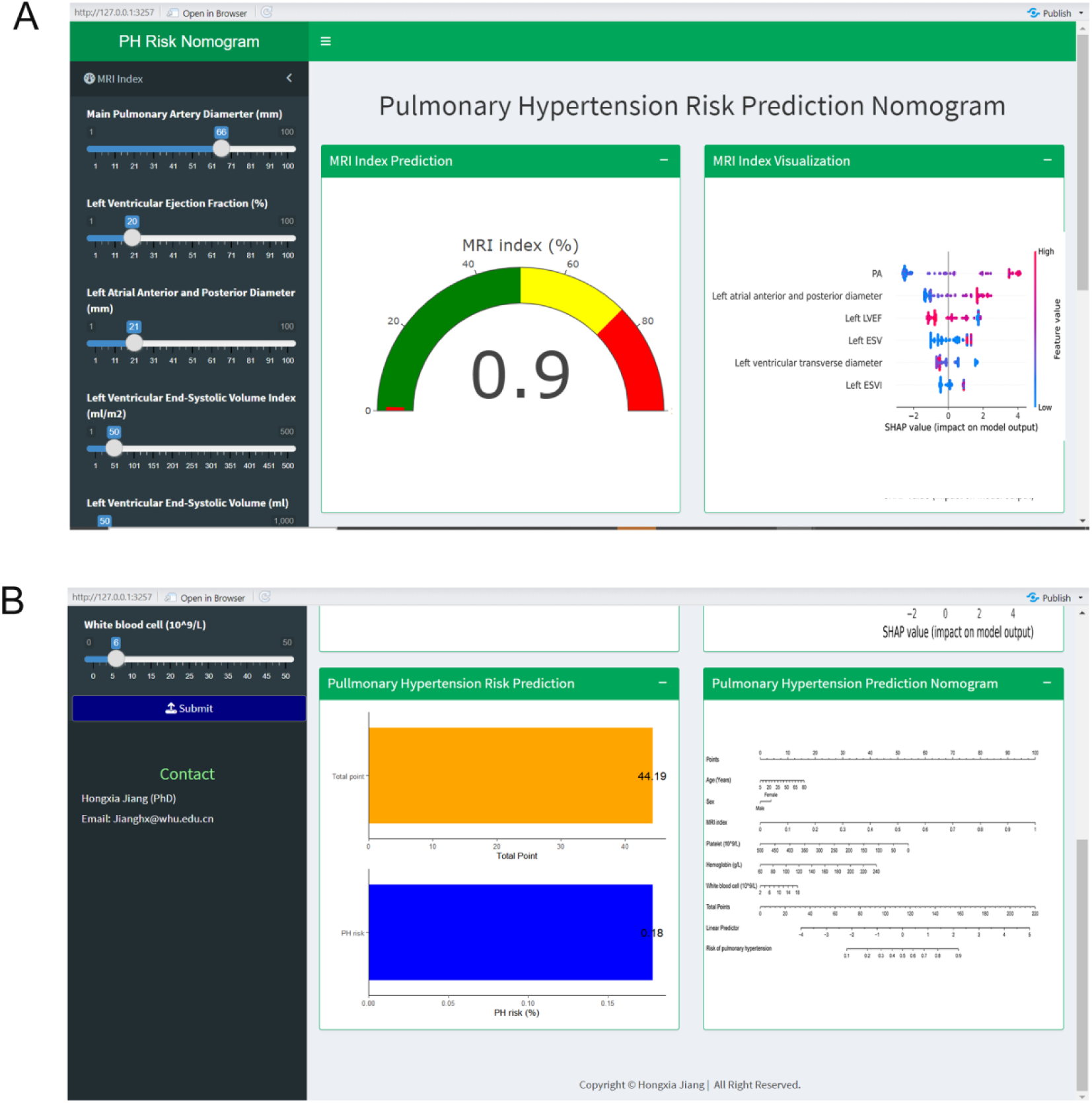
Risk prediction tools for pulmonary hypertension. (A) MRI-based pulmonary hypertension risk calculator presenting the index prediction and SHAP analysis, which explains the contribution of each feature to the model’s output.(B) Pulmonary hypertension risk prediction nomogram, utilizing clinical variables to calculate a total score, which corresponds to the estimated risk percentage of PH.

Figure 5B features a predictive nomogram based on clinical variables. The nomogram visually quantifies the contribution of each factor, and the cumulative score translates into a precise risk probability for PH. This tool streamlines the evaluation process, enabling practitioners to make swift and accurate risk assessments by integrating MRI and clinical features.

These tools enhance diagnostic precision by combining quantitative MRI measurements with critical clinical parameters, fostering a deeper understanding of PH risk and aiding in timely clinical decision-making. This is the URL of the prediction tool:https://jianghx.shinyapps.io/PH_Predictor/.

## Discussion

This study developed and validated a novel, multidimensional diagnostic model for PH that integrates MRI-derived features with clinical variables through advanced machine learning techniques. The results indicate significant improvements in predictive performance, interpretability, and clinical utility compared to traditional diagnostic methods. By integrating structural cardiac parameters and biochemical markers, this approach offers a comprehensive and accurate tool for early PH diagnosis, advancing the precision medicine paradigm.

A notable highlight of our study is that all cases in the PH group were verified by right heart catheterization, with an average pulmonary artery pressure >20 mmHg, and had complete cardiac MRI data. All individuals in the control group also had complete cardiac MRI data, with all cardiac function parameters within normal ranges. In other words, Our case group was diagnosed with PH by the gold standard, while our control group was confirmed by cardiac MRI to have no structural or functional abnormalities in the heart, ensuring there are no false positives or false negatives. Although these stringent inclusion criteria limited the number of cases in our study, they greatly enhanced the accuracy and clinical applicability of our data and model.

Our analysis identified several critical MRI-based predictors, PA diameter, left atrial anterior and posterior diameter, and left ventricular ejection fraction which are pathophysiologically significant in PH. These parameters were selected using RFE and validated through 10-fold cross-validation, ensuring their robustness in predicting PH. The enlargement of RA and LA reflects increased pulmonary circulation pressure and heightened right ventricular load, hallmark features of PH[8]. Similarly, an enlarged PA diameter is indicative of elevated pulmonary vascular resistance, directly correlating with the disease’s progression[9].

The inclusion of clinical variables such as white blood cell count, hemoglobin levels, and platelet count further enhanced the model’s predictive power[10,11]. Elevated white blood cell count, often associated with inflammation, reflects the inflammatory processes implicated in the progression of PH. Reduced hemoglobin levels may lead to impaired oxygen delivery, exacerbating the symptoms of PH. Abnormal platelet counts, particularly increased levels, are indicative of the hypercoagulable state frequently observed in PH patients. Integrating these hematological biomarkers with MRI-derived structural metrics significantly improved the model’s diagnostic accuracy and interpretability.

Machine learning algorithms, including XGBoost, Random Forest, and Logistic Regression, were evaluated for their predictive performance[12–15]. XGBoost consistently outperformed the other models, achieving an AUC of 1. Its ability to handle high-dimensional data and model complex, nonlinear relationships makes it particularly suitable for this application. Furthermore, the integration of SHAP analysis provided critical interpretability to the XGBoost model, highlighting the relative importance of key variables like LA size and PA diameter[16]. This transparency bridges the gap between complex machine learning models and their practical application in clinical settings.

Traditional diagnostic approaches for PH, including right heart catheterization and echocardiography, have notable limitations[17]. Catheterization, while considered the gold standard, is invasive and associated with procedural risks. Echocardiography, on the other hand, relies on tricuspid regurgitation velocity to estimate pulmonary artery pressure but is prone to operator-dependent variability and inaccuracies.

Recent advances in machine learning and biomarker discovery have sought to improve PH diagnostics. Studies by Boucly et al. and Hirata et al. explored the use of cytokines and echocardiographic parameters for PH risk prediction, respectively[18,19]. While these approaches demonstrated improved predictive accuracy, they relied on single-modality data and lacked the multidimensional integration seen in our study. Furthermore, prior studies often employed traditional statistical models or basic machine learning methods, which may fail to capture the intricate interactions between variables[20]. Our research addresses these gaps by combining MRI-derived features with clinical biomarkers, utilizing advanced feature selection methods like RFE, and implementing SHAP analysis for interpretability.

This comprehensive approach not only enhances predictive accuracy but also provides actionable insights into the pathophysiology of PH. The nomogram we developed offers a user-friendly interface for risk calculation, translating complex statistical outputs into clinically relevant formats. Compared to previous models, our framework demonstrates superior robustness and clinical utility.

The diagnostic tools developed in this study offer significant clinical benefits. The MRI-based risk calculator provides a non-invasive method to assess structural cardiac abnormalities, facilitating early detection of high-risk patients. This is particularly important in PH, where timely intervention can significantly improve outcomes. The nomogram further simplifies risk assessment by enabling clinicians to calculate individual risk scores based on both MRI-derived and clinical variables, promoting personalized treatment strategies.

By integrating SHAP analysis, our models provide transparency, allowing clinicians to understand the contributions of individual features to risk predictions. This interpretability builds trust in the model’s outputs and supports informed clinical decision-making. The combination of high accuracy, user-friendly tools, and actionable insights makes our framework a valuable addition to the current diagnostic arsenal for PH.

However, despite these strengths, our study is not without limitations. First, the sample size was relatively small and derived from a single center, which may limit the generalizability of the findings. Although rigorous cross-validation was employed to mitigate this limitation, future research should aim to validate the model in larger, multicenter datasets to ensure its external validity. Second, while MRI provides unparalleled structural detail, its cost and accessibility may hinder its widespread adoption in routine clinical practice. Exploring the integration of more commonly available imaging modalities, such as echocardiography, could improve the model’s practicality. Finally, our study utilized static data for analysis, and dynamic changes in biomarkers or imaging parameters over time were not considered. Incorporating longitudinal data in future studies could provide deeper insights into disease progression and further enhance the predictive accuracy of the model.

Future research should focus on validating the model in larger, diverse populations, incorporating dynamic and emerging biomarkers, and automating workflows to enhance clinical adoption and expand its utility to other diseases.

## Conclusion

This study presents a groundbreaking diagnostic framework for PH, integrating MRI-derived features and clinical variables through advanced machine-learning techniques. The model exhibits exceptional predictive accuracy, interpretability, and clinical utility, paving the way for improved early diagnosis and personalized treatment strategies. By addressing the limitations of traditional diagnostic methods and incorporating cutting-edge technologies, our approach represents a significant advancement in the field of PH diagnostics and holds the potential to transform clinical practice.

## Founding

This study was funded by the following programs: The National Natural Science Foundation of China (No.82300078), the Fundamental Research Funds for the Central Universities (No.2042023kf0059), Wuhan University Clinical Medicine + Youth Supporting Program (No.413000557), the Climbing Project for Medical Talent of Zhongnan Hospital of Wuhan University (Grant No. PDJH202406), Science and Technology Innovation Cultivation Funding of Zhongnan Hospital of Wuhan University (No. CXPY2023060).

## Data Availability

All the original data involved in this investigation are available on reasonable request to the first author or corresponding author.

---

**Table 1.**
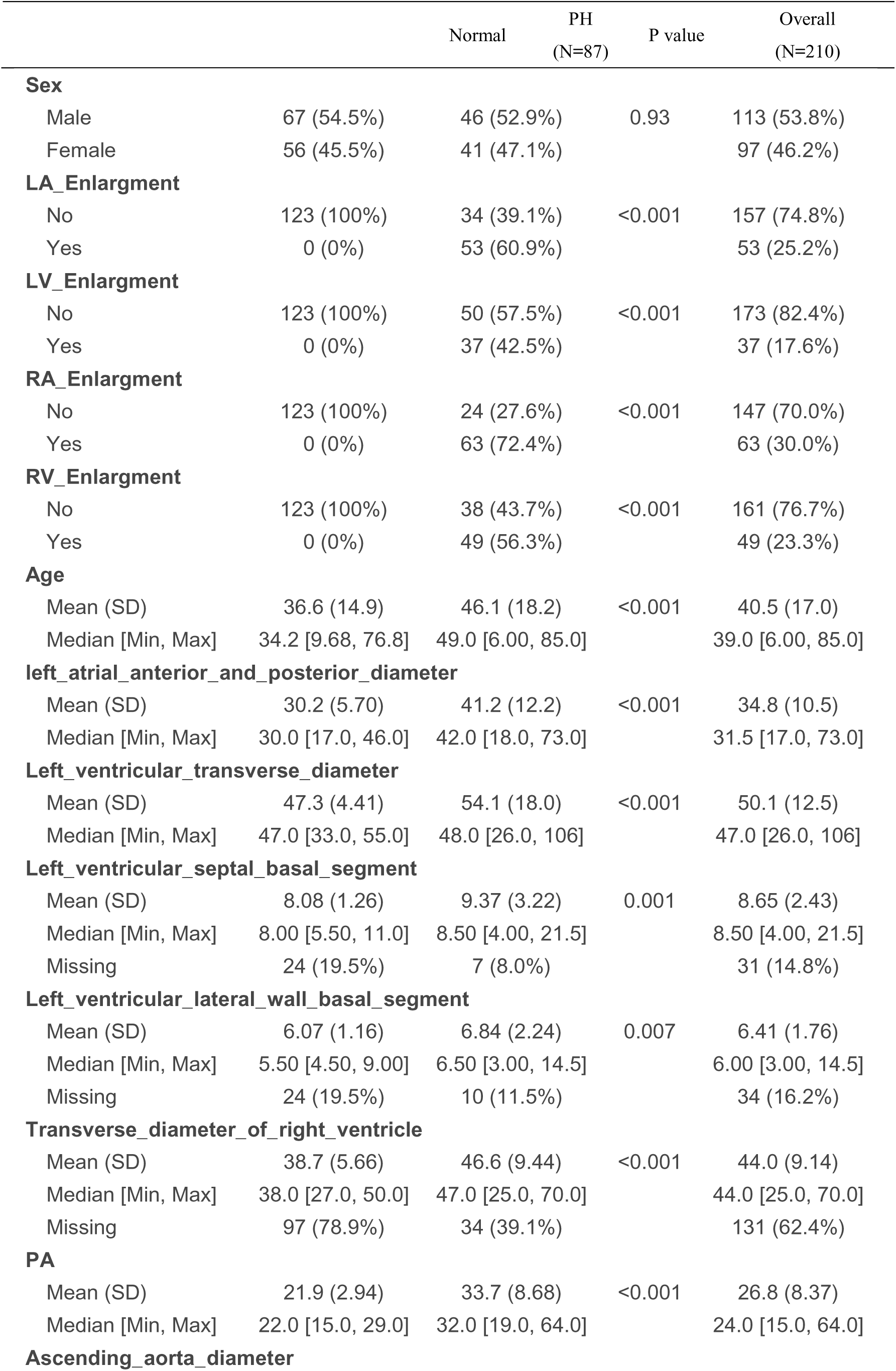
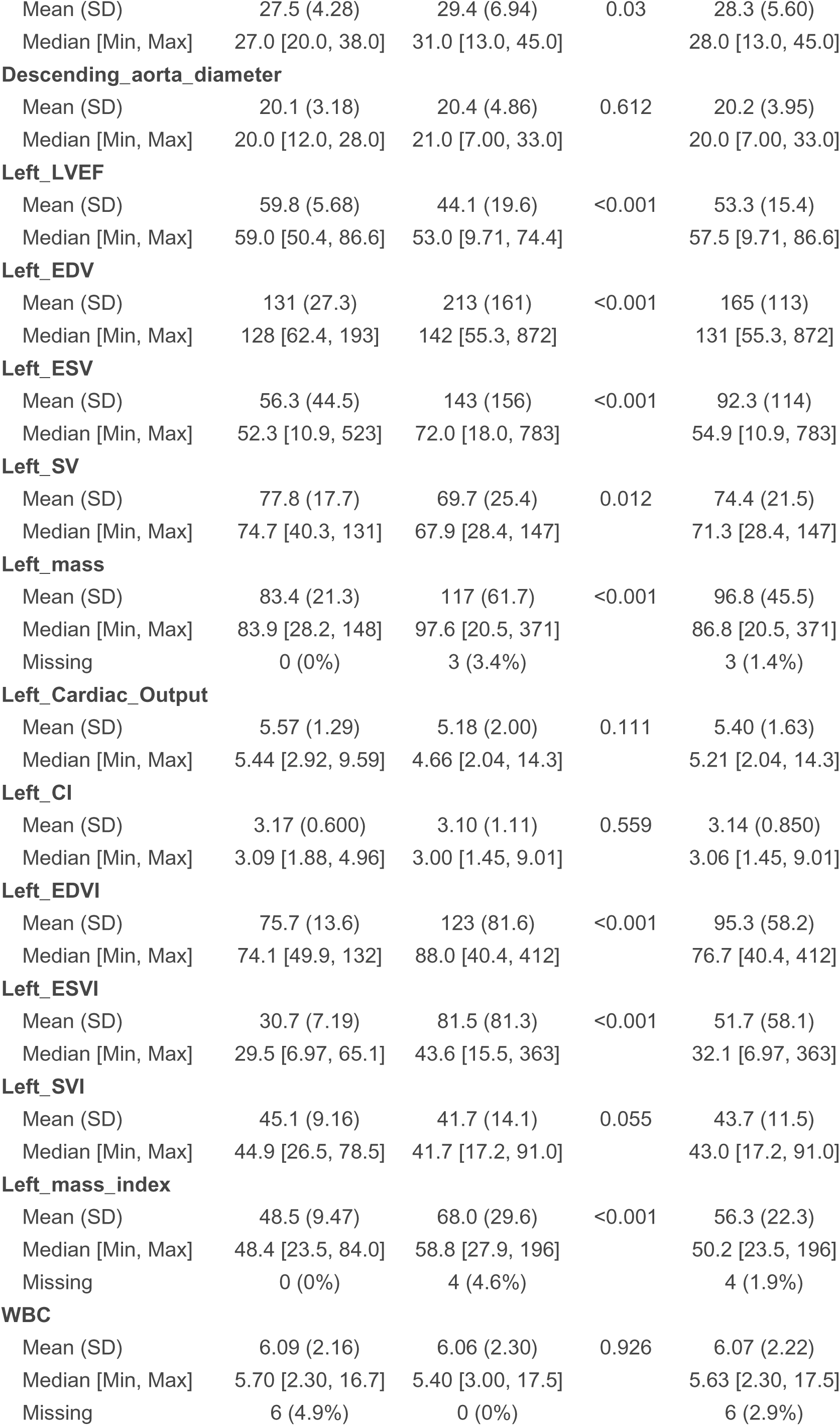
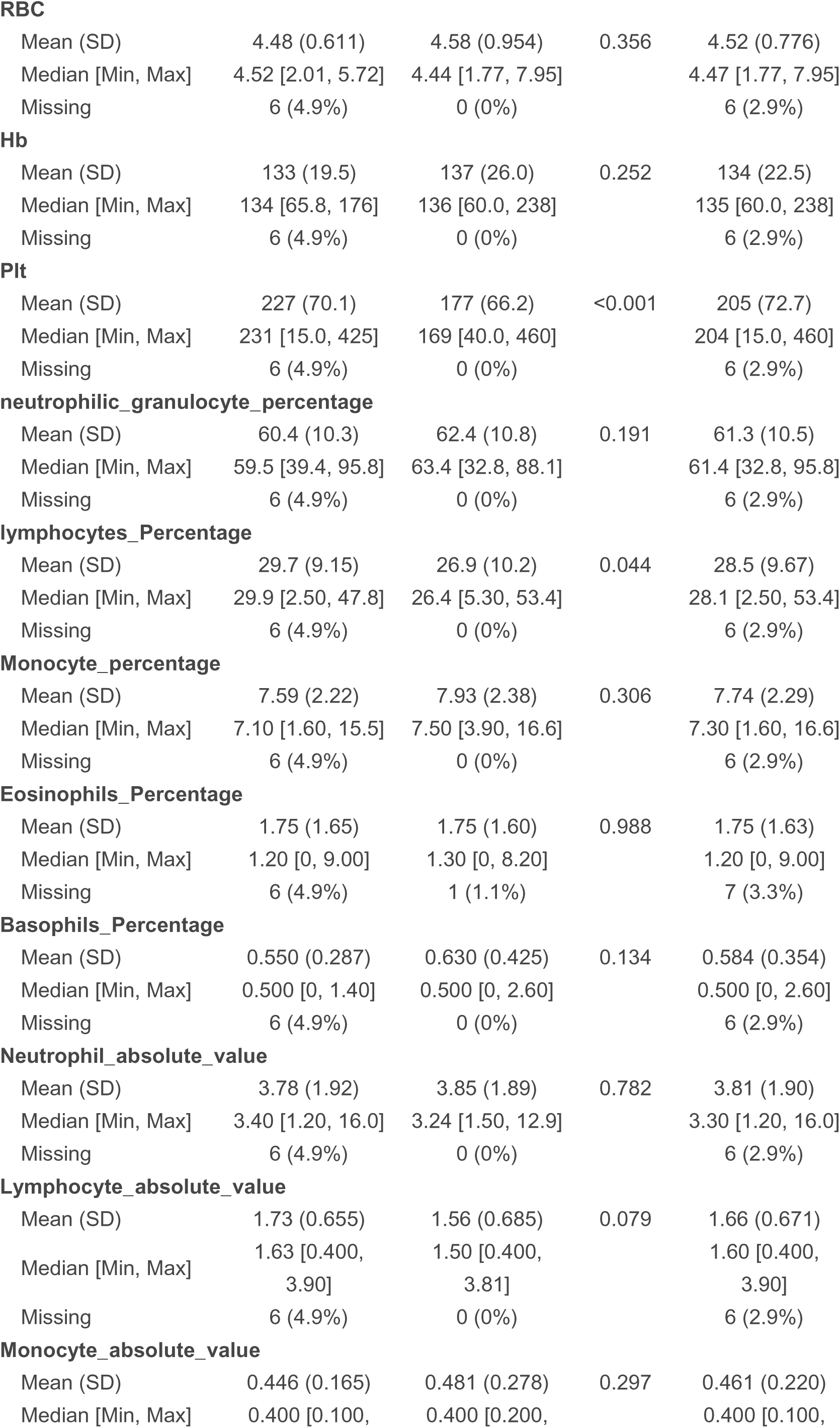
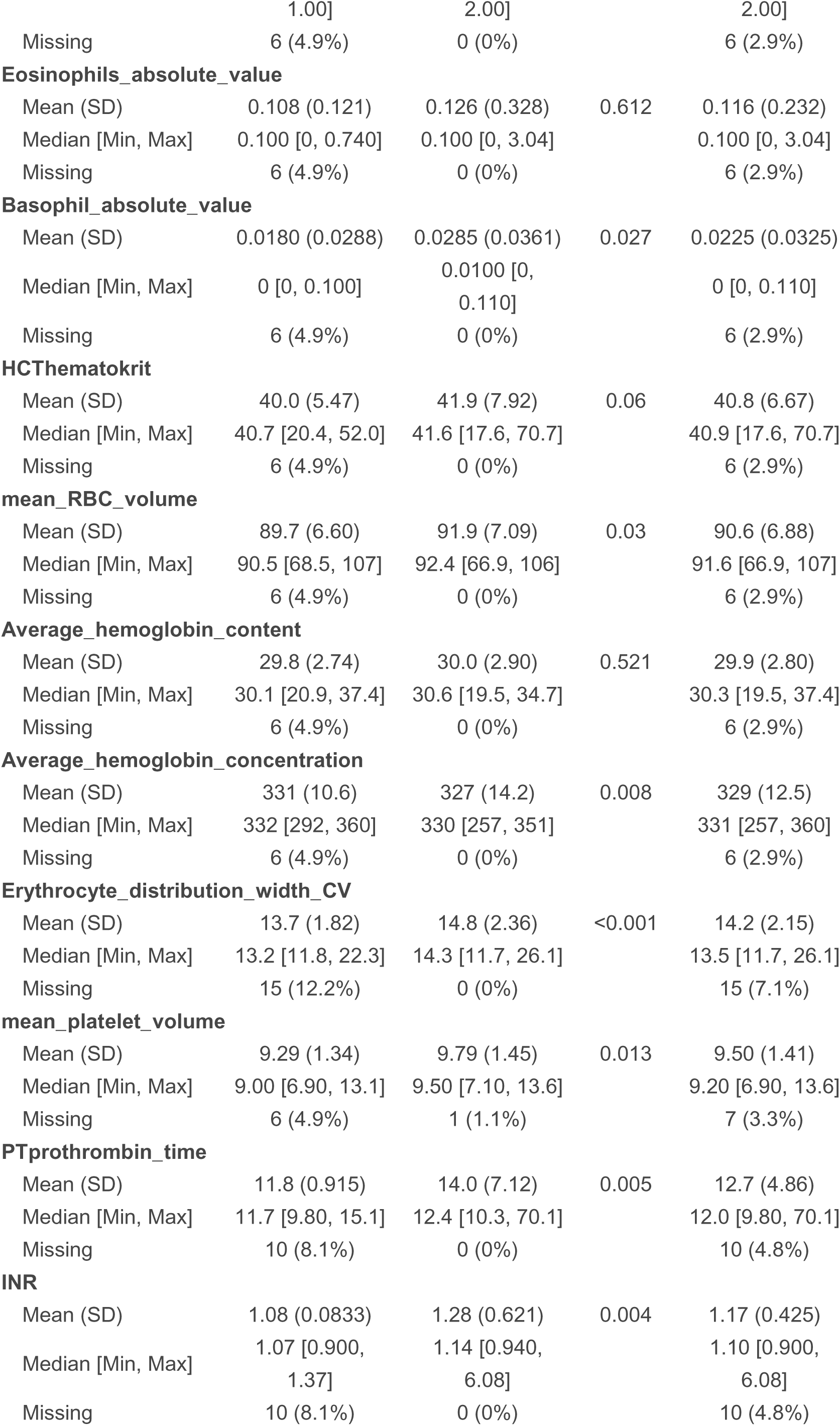
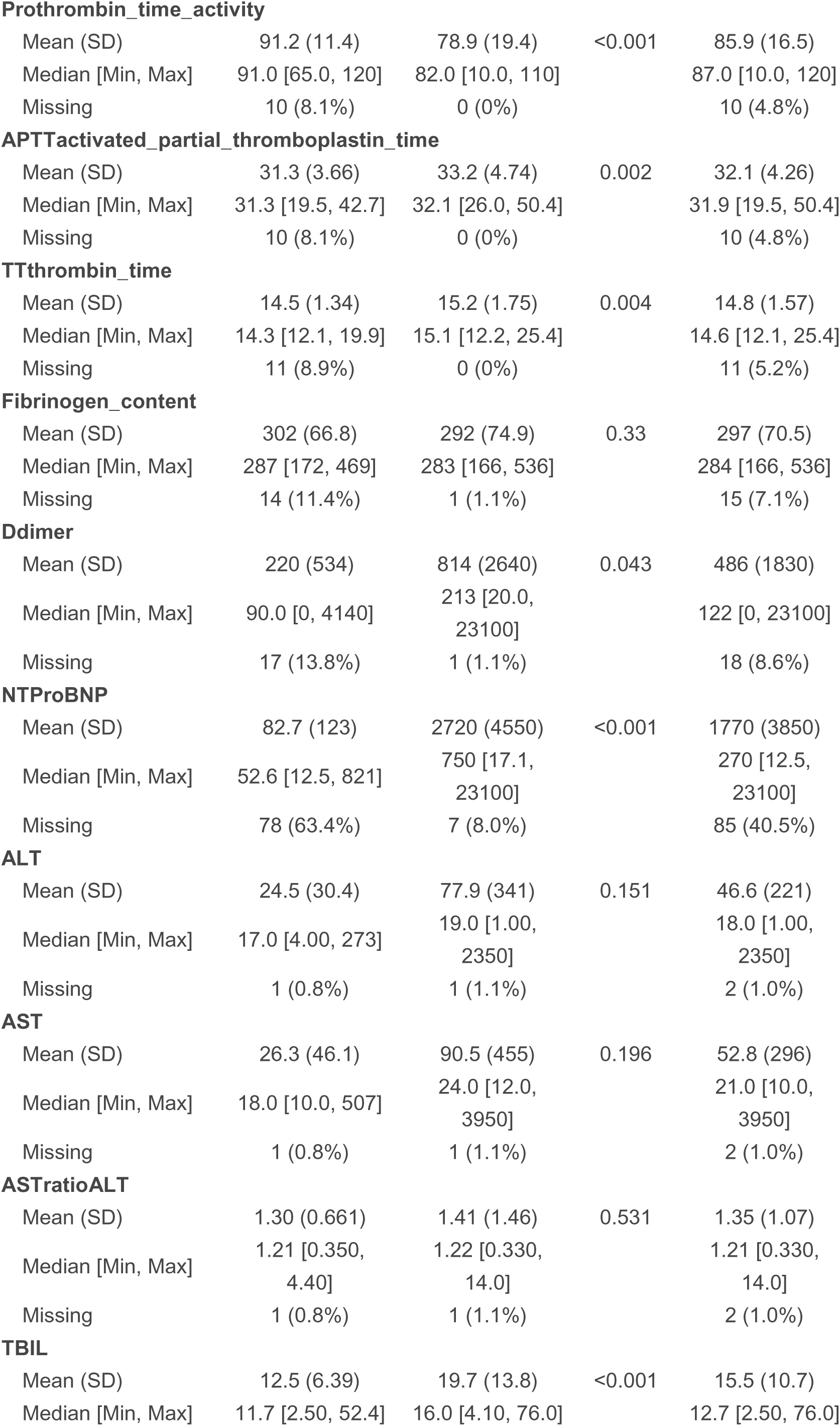
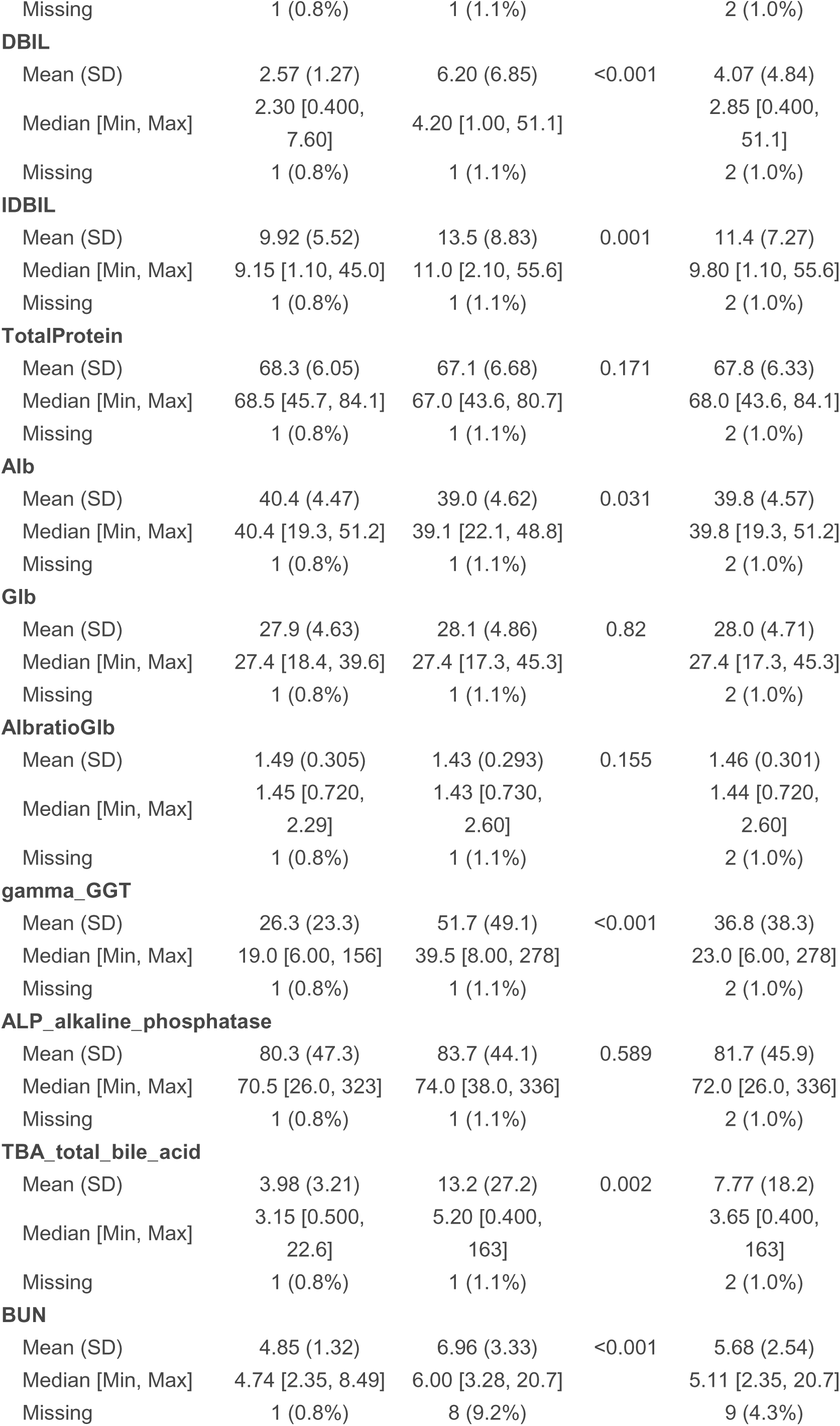
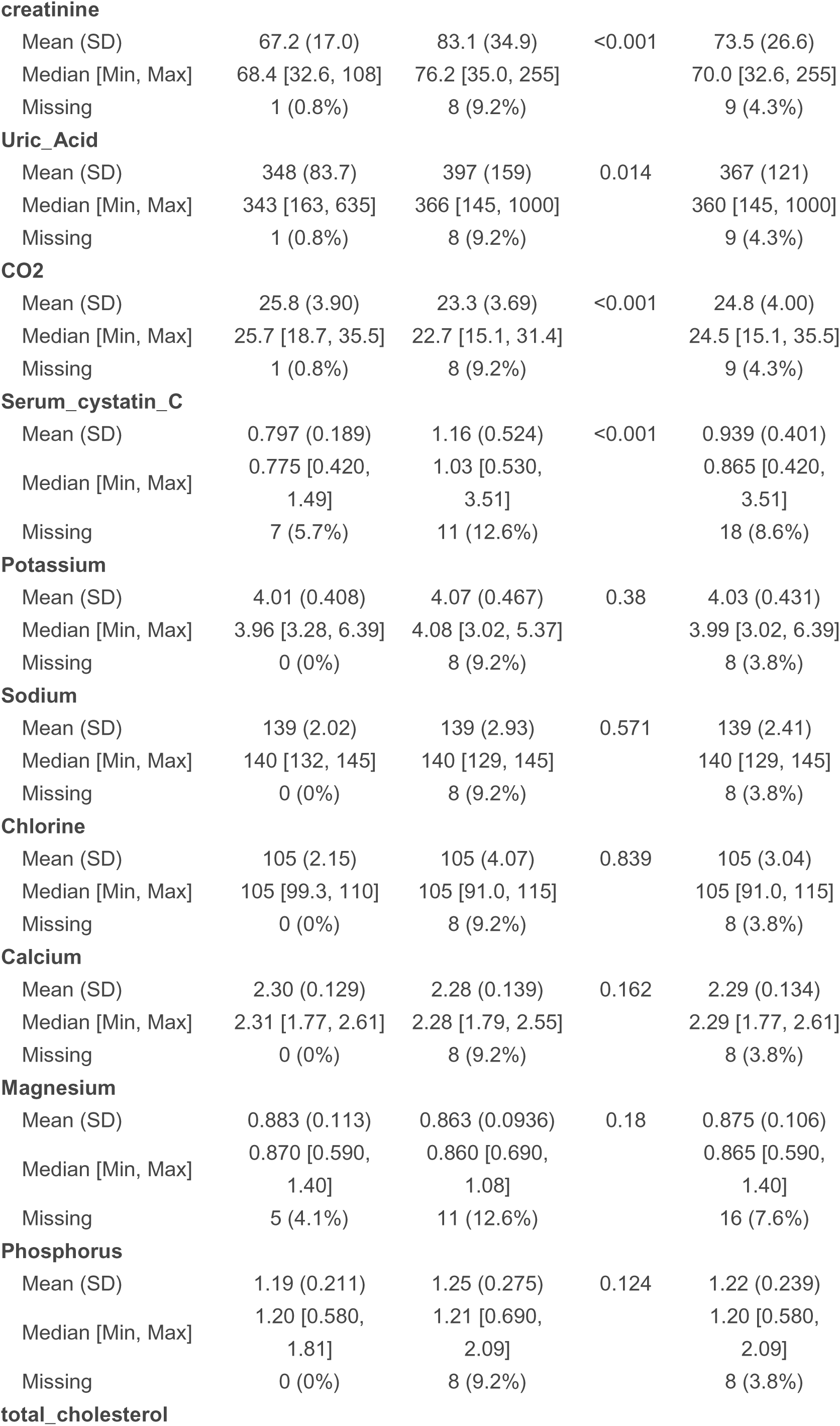
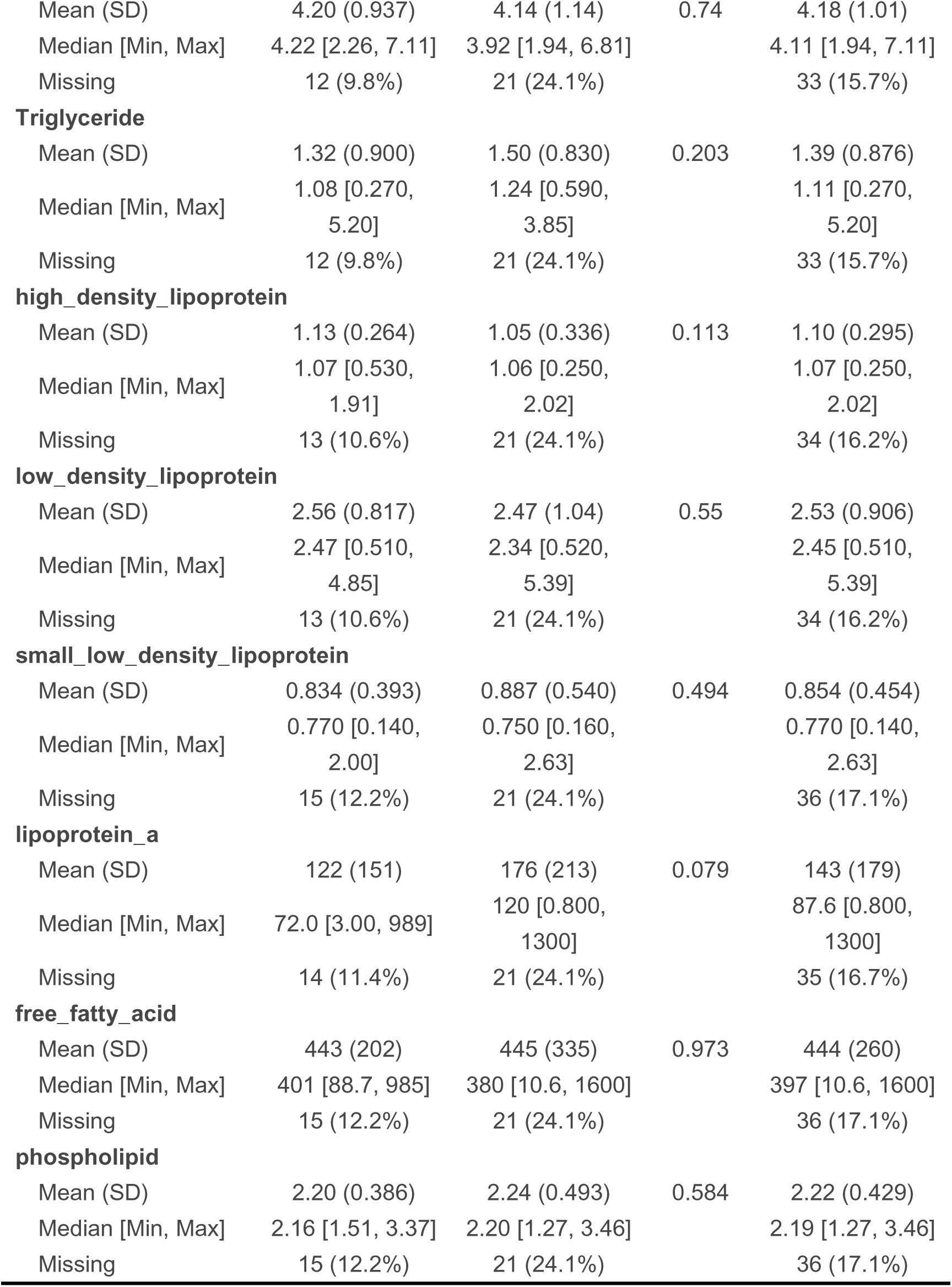

